# Characterization of e-cigarette use among school-attending adolescents in Thailand

**DOI:** 10.64898/2026.11.26.26369108

**Authors:** Chunxia He, Wit Wichaidit

## Abstract

**Background:** The objective of this study is to describe the prevalence and characteristics of e-cigarette use among school-going adolescents in Thailand, disaggregated by sex assigned at birth and year level.

**Methods:** We conducted a school-based cross-sectional study by requesting students in Matthayom 1 (Year 7), Matthayom 3 (Year 9), and Matthayom 5 or Vocational Certificate 2 (Year 11) at randomly selected schools and classrooms to complete self-administered questionnaire with a section on e-cigarette use. Data were analyzed using descriptive statistics with adjustment for sampling weight and survey design.

**Results:** A total of 23,931 students answered the study questionnaires with adequately complete responses. The weighted prevalence of e-cigarette ever use was 22.3%, and the prevalence of current use was 7.8%. Among the current users, 31.1% reported moderate to severe addiction. The most common method of obtaining e-cigarette was by own purchase (45.2%), and electric pods were the most common type of e-cigarette (66.6%). Differences by sex were less notable compared to differences by educational system: students in the vocational education system had significantly higher prevalence of ever using e-cigarette than students in the general education system.

**Discussion and Conclusion:** Our study results offer potentially useful basic information for stakeholders in tobacco control and adolescent health. However, limitations regarding potential biases, lack of key details on e-cigarette use behaviors, and the inability to generalize the findings to adolescents not enrolled in formal education should be considered as caveats in the interpretation of the study findings.

## INTRODUCTION

Globally, the use of electronic cigarettes (e-cigarettes or “vapes”) has become a public health concern. In 2025, the World Health Organization (WHO) estimated that more than 100 million people worldwide used e-cigarettes. In countries with available data, adolescents were approximately nine times as likely as adults to use e-cigarettes(World Health Organization, 2025). The WHO has warned that e-cigarettes, often marketed as harm-reduction products, may contribute to a new generation of nicotine dependence, particularly among adolescents(World Health Organization, 2025). Previous research among Chinese university students found substantial gaps in awareness of vaping-related health risks and regulations, alongside nicotine dependence among users(He et al., 2026). Recent systematic reviews have linked e-cigarette use to adverse respiratory outcomes, including asthma and bronchitic symptoms, as well as cardiovascular effects(Golder et al., 2025; Kaur et al., 2025; Kundu et al., 2025). Moreover, nicotine exposure during adolescence may adversely affect brain development, raising concerns about the long-term health consequences of early vaping initiation(Reynolds et al., 2025).

The prevalence, patterns and associated factors of vaping among adolescents vary across countries, with differences in demographic, social, and regulatory contexts potentially contributing to this variation(Ko et al., 2024). For example, the prevalence of current vaping was 3.0% in China(Qi et al., 2022) and 6.1% in Vietnam(Phan et al., 2025) to 14.9% in Malaysia(Mohd Yusoff et al., 2026) and 7.8% among US(Park-Lee et al., 2024) high school students. Thailand presents a distinctive regulatory context, where e-cigarette imports and sales have been prohibited since 2014-2015(Patanavanich & Glantz, 2021), while cannabis was decriminalized in 2022(Charoenwisedsil et al., 2023). Previous studies have reported positive associations between e-cigarette use and cannabis use among adolescents(Golder et al., 2025; Phetphum et al., 2026). Despite restrictions on e-cigarettes, e-cigarette use is common among Thai adolescents, with reported prevalence estimates of 3.7% in 2019(Patanavanich et al., 2021), 9.6 % in 2023(Phetphum et al., 2024), and 5.3% in 2024-2025(Patanavanich et al., 2026). Given the variation in e-cigarette use by sex and age(Patanavanich et al., 2026), continued monitoring of vaping behaviors among Thai adolescents with disaggregation by sex and year level (as a proxy for age and educational system) is needed in order to obtain timely empirical evidence for stakeholders in tobacco control and adolescent health. The objective of this study is to describe the prevalence and characteristics of e-cigarette use among school-going adolescents in Thailand, disaggregated by sex assigned at birth and year level.

## METHODS

### Study design and setting

We analyzed data from a nationally-representative cross-sectional study, officially titled the National School Surveillances of Alcohol, Tobacco, Substance Use, and Health-Risk Behaviors among Secondary Students or Equivalent (henceforward, "the Survey"). Data collection for the Survey took place from July to November, 2024.

### Study population

The study population comprised Mathayom 1 (Grade 7), Mathayom 3 (Grade 9), and Mathayom 5 (Grade 11) students in general secondary schools, and second-year vocational certificate students (equivalent to Grade 11), divided into either the commercial stream or the technical stream. These grades were selected to cover a broad adolescent age range (approximately 12-17 years) with minimal age overlap.

### Study participants and sample size calculation

Study participants included currently enrolled general education students in Matthayom 1 (Year 7), Matthayom 3 (Year 9), and Matthayom 5 (Year 11) levels, or students in the Vocational Certificate Year 2 (Year 11) level in the selected schools who were present in the selected classrooms at the time of the Survey. The investigators excluded students who had difficulty reading and completing the questionnaires and those under suspension from the study. The sample size was calculated based on the main objective of monitoring changing trends in risk behaviors, based on the prevalence of alcohol consumption during the students’ lifetime. Sample size calculation was made with the assumption that the prevalence would be equal to that of a previous study(Assanangkornchai et al., 2018) at 34 percent (p=0.34) with a margin of error of no greater than 2 percent (delta=0.02) at 95% level of confidence. The investigator then adjusted for the design effect of 2.0, and expanded the sample size by a factor of 5 to enable estimation of the prevalence in all five study regions, and assumed 10 percent refusal to participate or complete the questionnaire, yielding the final sample size of 23,705 students, with the estimated range between 23,900 to 30,400 students from 109 schools across 21 provinces and Bangkok.

### Study instrument

The survey used an anonymous, self-administered questionnaire adapted from the fourth and fifth rounds of the Thai national school-based surveillance survey(Assanangkornchai et al., 2018), which drew on the Monitoring the Future questionnaire(Johnston et al., 2014) and relevant literature. The questionnaire was reviewed by experts in adolescent mental health and substance use and pilot-tested among 30 students to assess its reliability. This study used items on student characteristics (Section A), family, school, and community environment (Section B), and cigarette smoking and e-cigarette use (Section D). Cigarette nicotine dependence was assessed using the Fagerström Test for Nicotine Dependence(Heatherton et al., 1991), while e-cigarette dependence was assessed using a school-adapted version of the Penn State Electronic Cigarette Dependence Index(Foulds et al., 2015). The questionnaire took approximately 30-50 minutes to complete.

### Study variables

#### Outcome (e-cigarette use behaviors)

The investigators designed questions to measure electronic cigarette use based on the questions used in a previous round of the Survey(Wichaidit et al., 2024). Questions included lifetime e-cigarette use, latest occasion of e-cigarette use, source of e-cigarette in the past 12 months, type of e-cigarette used in the past 12 months (with accompanying images), as well as an e-cigarette addiction screening test based on the Penn State Electronic Cigarette Dependence Index(Foulds et al., 2015). The investigators used the same scoring system and cut-off values for categorization as the original instrument: 1) Not addicted (0 to 3 points); 2) Mildly addicted (4 to 8 points); 3) Moderately addicted (9 to 12 points); 4) Severely addicted (13 points or higher).

#### Disaggregating variables (participant characteristics)

In the first section of the questionnaire, investigators asked the student to self-report their sex assigned at birth, religion, and the amount of allowance per week (in Thai Baht or THB). The data entry team entered information regarding the level of study of the students by default.

### Data collection

The investigators selected the participants using a multistage stratified cluster sampling method. A total of 21 provinces in all regions of Thailand were randomly selected, and Bangkok was purposively selected. In each selected province, three general secondary schools (urban public, rural public, and private) and two vocational schools (commercial and technical) were randomly selected. In Bangkok, two general secondary schools and two vocational schools were selected. Classrooms were randomly selected from the eligible grades, while all classrooms were included in smaller schools.

Researchers randomly selected provinces and obtained updated lists of schools. Schools were then selected using simple random sampling. Research staff contacted the selected schools, collected basic school information, and randomly selected classrooms. Students were informed about the study, and group consent was obtained before they voluntarily completed the questionnaires. No personal identifying information was collected to protect participants’ confidentiality.

### Data management

All questionnaires were checked for accuracy and completeness by research staff upon collection. Data from paper questionnaires were entered using the KoboToolbox platform. Blank questionnaires were considered refusals and excluded. After data entry, questionnaires with more than 30% of all questions that were asked to all students (i.e., not under skip patterns) being unanswered were considered incomplete and excluded from the analyses. The investigators then cleaned the remaining data for analyses.

### Data Analysis

In the clean data set, the investigators imputed the values of "Refuse to answer" to all responses that were left blank, except for allowance, in which the value of 50,000 THB per week or higher was treated as outliers to avoid over-skewing the weighted mean. The investigators then analyzed data using descriptive statistics with adjustment for sampling weight and the complex study design using the *survey* package in R. The investigators then disaggregated e-cigarette use behaviors by sex and by year of study.

### Ethical Considerations

We received Ethical Approval from the Human Research Ethics Unit of the Faculty of Medicine, Prince of Songkla University (approval number: REC.66-437-18-2). We also received a waiver of written informed consent from the Committee. The study complied with relevant national regulations and institutional policies and followed the principles of the Declaration of Helsinki.

## RESULTS

A total of 24,341 participants returned the questionnaire, 23,931 of whom (98.3%) provided complete responses and were included in the analysis data. The proportion of participants who were male and female was approximately equal (*Table 1*). Most participants were Buddhist and received slightly more than 500 THB per week in allowance. Nearly one quarter (22.3%) of the participants had used e-cigarettes in their lifetime, although only 7.8% were current users. Among participants who had used e-cigarettes in the past 12 months, the most common way to obtain e-cigarettes was the participants’ own purchase (45.2%), and electric pods were the most common type of e-cigarette (66.6%). Among the current users, nearly one-third (31.1%) reported moderate to severe addiction.

**Table 1.** General characteristics and e-cigarette use behaviors of the study participants (n=23,931 participants, unless otherwise indicated)

| Characteristic | Percentage ± Standard Error (SE) |
|---|---|
| **Sex assigned at birth** |  |
| Male | 47.9% ± 1.4% |
| Female | 49.5% ± 1.4% |
| Intersex | 0.8% ± 0.1% |
| Refused to answer | 1.8% ± 0.2% |
| **Level of study** |  |
| Matthayom 1 (Year 7) | 34.6% ± 1.2% |
| Matthayom 3 (Year 9) | 33.7% ± 1.0% |
| Matthayom 5 (Year 11, general education) | 21.9% ± 0.7% |
| Vocational Certificate 2 (Year 11, vocational) - Commercial track | 4.3% ± 1.1% |
| Vocational Certificate 2 (Year 11, vocational) - Industrial technology track | 5.3% ± 1.2% |
| Refused to answer | 0.1% ± 0.0% |
| **Religion** |  |
| Buddhism | 92.3% ± 1.5% |
| Islam | 4.6% ± 1.4% |
| Christianity | 2.1% ± 0.6% |
| Others | 0.5% ± 0.1% |
| Refused to answer | 0.4% ± 0.1% |
| **Amount of allowance per week (THB)** (weighted mean ± standard error) | 549 ± 11 |
| **History of e-cigarette use** |  |
| Never used in lifetime | 77.7% ± 1.2% |
| Former users, long-time quitters (used in lifetime but not in the past 12 months) | 7.4% ± 0.4% |
| Former users, recent quitters (used in the past 12 months but not in the past 30 days) | 4.0% ± 0.2% |
| Current users (used in the past 30 days) | 7.8% ± 0.6% |
| Incomplete information or refused to answer | 3.1% ± 0.3% |
| **Among recent quitters and current users: Most common source of e-cigarette in the past 12 months** | (n=3,261) |
| Own purchase | 45.2% ± 1.1% |
| Nominee purchase (buyer age more than 20 years) | 4.0% ± 0.4% |
| Nominee purchase (buyer age less than 20 years) | 3.8% ± 0.4% |
| Given by others (buyer age more than 20 years) | 4.2% ± 0.5% |
| Given by others (buyer age less than 20 years) | 6.0% ± 0.6% |
| Stolen | 0.8% ± 0.2% |
| Others | 8.5% ± 0.7% |
| Refused to answer | 27.4% ± 1.1% |
| **Among recent quitters and current users: Type of e-cigarette used in the past 12 months** | (n=3,261) |
| Type 1 (looking similar to cigarettes) | 6.2% ± 0.5% |
| Type 2 (pen) | 0.5% ± 0.1% |
| Type 3 (tank) | 4.4% ± 0.4% |
| Type 4 (electric pods) | 66.6% ± 1.4% |
| Type 5 (toy-like pods) | 9.1% ± 0.8% |
| Others | 3.1% ± 0.4% |
| Refused to answer | 10.0% ± 0.7% |
| **E-cigarette addiction screening test results (among current users only)** | (n=2,248) |
| Not addicted (0-3 points) | 25.2% ± 1.1% |
| Mildly addicted (4-8 points) | 29.4% ± 1.3% |
| Moderately addicted (9-12 points) | 18.1% ± 0.8% |
| Severely addicted (13 points or higher) | 13.0% ± 0.8% |
| Incomplete information or refused to answer | 14.2% ± 1.3% |

When disaggregated by sex (*Table 2*), the participants had similar histories of e-cigarette use, source of e-cigarette, type of e-cigarette, and addiction screening test results. However, when disaggregated by year level and school system, there were notable differences (*Table 3*). Students in Vocational Certificate Program Year 2 were more likely to be current e-cigarette users, particularly those in the Technical stream. Younger students (those in Matthayom 1, or Year 7) were less likely to report having purchased the e-cigarette by themselves. Students in vocational certificate programs were more likely to report having used e-cigarettes in the form of toy-like objects. However, there were no notable differences in the distribution of e-cigarette addiction by year level and school systems.

**Table 2.**
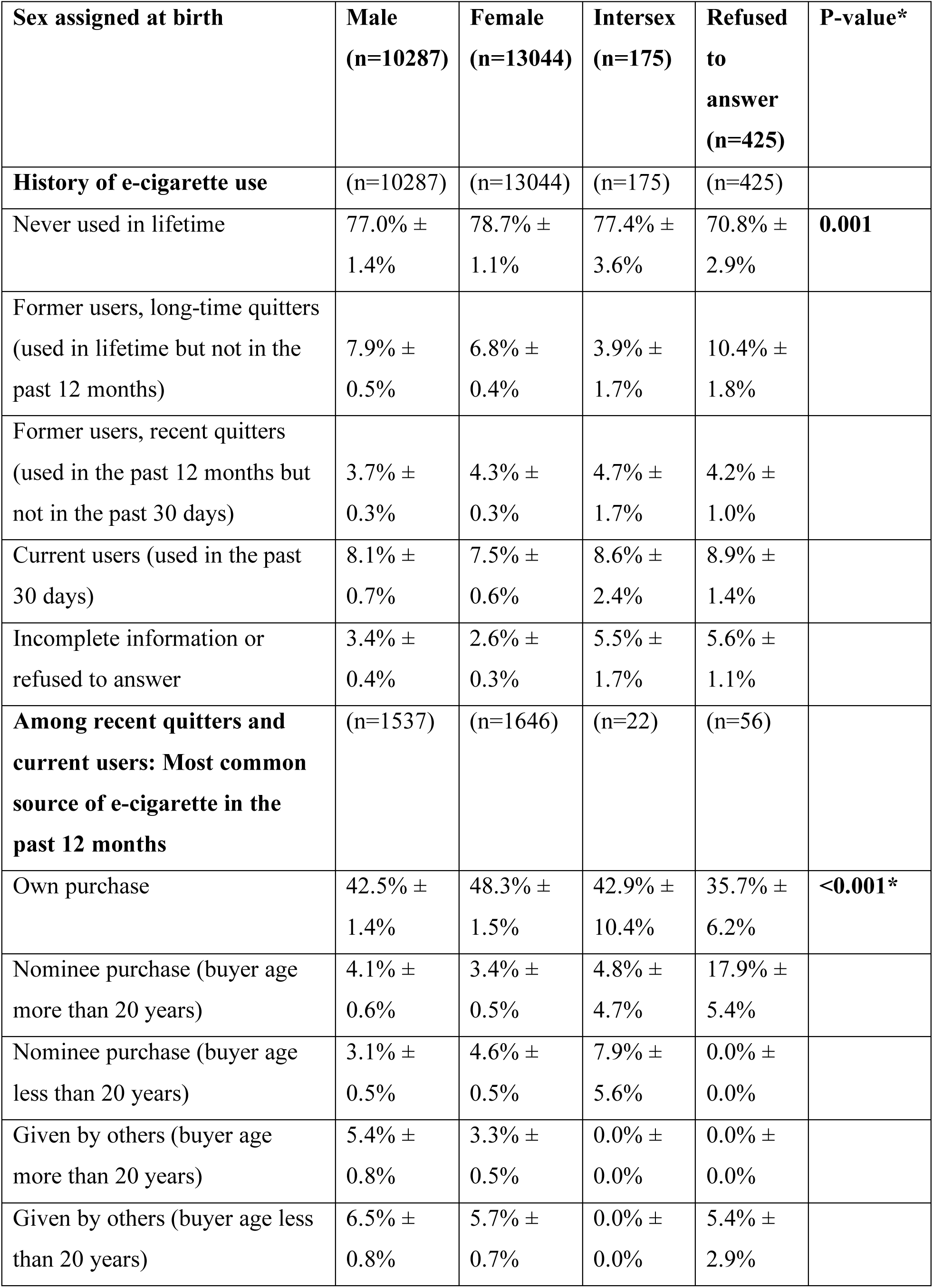
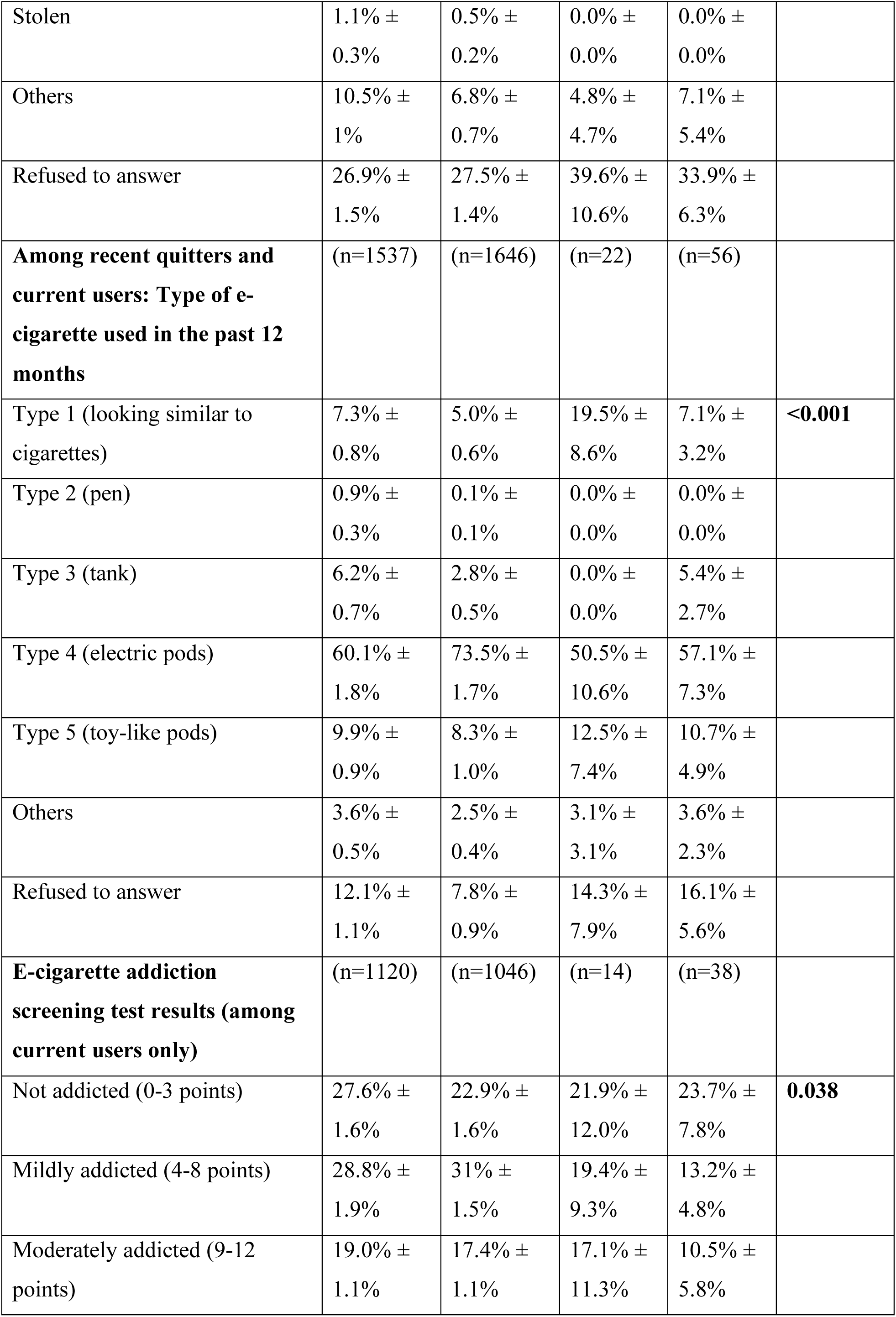
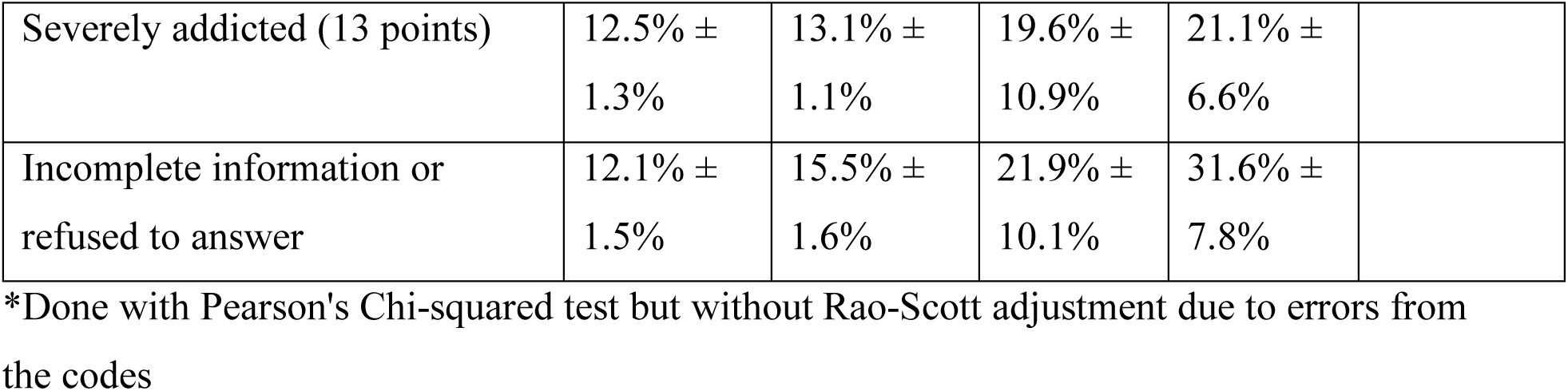
E-cigarette use behaviors among the study participants, disaggregated by sex assigned at birth (Column percent; mean ± SE)

| **Sex assigned at birth** | **Male (n=10287)** | **Female (n=13044)** | **Intersex (n=175)** | **Refused to answer (n=425)** | **P-value\*** |
|---|---|---|---|---|---|
| **History of e-cigarette use** | (n=10287) | (n=13044) | (n=175) | (n=425) |  |
| Never used in lifetime | 77.0% ± 1.4% | 78.7% ± 1.1% | 77.4% ± 3.6% | 70.8% ± 2.9% | **0.001** |
| Former users, long-time quitters (used in lifetime but not in the past 12 months) | 7.9% ± 0.5% | 6.8% ± 0.4% | 3.9% ± 1.7% | 10.4% ± 1.8% |  |
| Former users, recent quitters (used in the past 12 months but not in the past 30 days) | 3.7% ± 0.3% | 4.3% ± 0.3% | 4.7% ± 1.7% | 4.2% ± 1.0% |  |
| Current users (used in the past 30 days) | 8.1% ± 0.7% | 7.5% ± 0.6% | 8.6% ± 2.4% | 8.9% ± 1.4% |  |
| Incomplete information or refused to answer | 3.4% ± 0.4% | 2.6% ± 0.3% | 5.5% ± 1.7% | 5.6% ± 1.1% |  |
| **Among recent quitters and current users: Most common source of e-cigarette in the past 12 months** | (n=1537) | (n=1646) | (n=22) | (n=56) |  |
| Own purchase | 42.5% ± 1.4% | 48.3% ± 1.5% | 42.9% ± 10.4% | 35.7% ± 6.2% | **<0.001\*** |
| Nominee purchase (buyer age more than 20 years) | 4.1% ± 0.6% | 3.4% ± 0.5% | 4.8% ± 4.7% | 17.9% ± 5.4% |  |
| Nominee purchase (buyer age less than 20 years) | 3.1% ± 0.5% | 4.6% ± 0.5% | 7.9% ± 5.6% | 0.0% ± 0.0% |  |
| Given by others (buyer age more than 20 years) | 5.4% ± 0.8% | 3.3% ± 0.5% | 0.0% ± 0.0% | 0.0% ± 0.0% |  |
| Given by others (buyer age less than 20 years) | 6.5% ± 0.8% | 5.7% ± 0.7% | 0.0% ± 0.0% | 5.4% ± 2.9% |  |
| Stolen | 1.1% ± 0.3% | 0.5% ± 0.2% | 0.0% ± 0.0% | 0.0% ± 0.0% |  |
| Others | 10.5% ± 1% | 6.8% ± 0.7% | 4.8% ± 4.7% | 7.1% ± 5.4% |  |
| Refused to answer | 26.9% ± 1.5% | 27.5% ± 1.4% | 39.6% ± 10.6% | 33.9% ± 6.3% |  |
| **Among recent quitters and current users: Type of e-cigarette used in the past 12 months** | (n=1537) | (n=1646) | (n=22) | (n=56) |  |
| Type 1 (looking similar to cigarettes) | 7.3% ± 0.8% | 5.0% ± 0.6% | 19.5% ± 8.6% | 7.1% ± 3.2% | **<0.001** |
| Type 2 (pen) | 0.9% ± 0.3% | 0.1% ± 0.1% | 0.0% ± 0.0% | 0.0% ± 0.0% |  |
| Type 3 (tank) | 6.2% ± 0.7% | 2.8% ± 0.5% | 0.0% ± 0.0% | 5.4% ± 2.7% |  |
| Type 4 (electric pods) | 60.1% ± 1.8% | 73.5% ± 1.7% | 50.5% ± 10.6% | 57.1% ± 7.3% |  |
| Type 5 (toy-like pods) | 9.9% ± 0.9% | 8.3% ± 1.0% | 12.5% ± 7.4% | 10.7% ± 4.9% |  |
| Others | 3.6% ± 0.5% | 2.5% ± 0.4% | 3.1% ± 3.1% | 3.6% ± 2.3% |  |
| Refused to answer | 12.1% ± 1.1% | 7.8% ± 0.9% | 14.3% ± 7.9% | 16.1% ± 5.6% |  |
| **E-cigarette addiction screening test results (among current users only)** | (n=1120) | (n=1046) | (n=14) | (n=38) |  |
| Not addicted (0-3 points) | 27.6% ± 1.6% | 22.9% ± 1.6% | 21.9% ± 12.0% | 23.7% ± 7.8% | **0.038** |
| Mildly addicted (4-8 points) | 28.8% ± 1.9% | 31% ± 1.5% | 19.4% ± 9.3% | 13.2% ± 4.8% |  |
| Moderately addicted (9-12 points) | 19.0% ± 1.1% | 17.4% ± 1.1% | 17.1% ± 11.3% | 10.5% ± 5.8% |  |
| Severely addicted (13 points) | 12.5% ± 1.3% | 13.1% ± 1.1% | 19.6% ± 10.9% | 21.1% ± 6.6% |  |
| Incomplete information or refused to answer | 12.1% ± 1.5% | 15.5% ± 1.6% | 21.9% ± 10.1% | 31.6% ± 7.8% |  |

\*Done with Pearson's Chi-squared test but without Rao-Scott adjustment due to errors from the codes

**Table 3.** E-cigarette use behaviors among the study participants, disaggregated by level of study.

| **Level of Study** | **Matthayom 1 (Year 7) (n=6578)** | **Matthayom 3 (Year 9) (n=6407)** | **Matthayom 5 (Year 11, general education) (n=6263)** | **VC2 Commercial (n=2084)** | **VC2 Technical (n=2572)** | **P-value\*** |
|---|---|---|---|---|---|---|
| **History of e-cigarette use** | (n=6578) | (n=6407) | (n=6263) | (n=2084) | (n=2572) |  |
| Never used in lifetime | 83.2% ± 1.3% | 76.1% ± 1.6% | 79.7% ± 1.3% | 67.5% ± 2.5% | 51.9% ± 2.7% | **<0.001** |
| Former users, long-time quitters (used in lifetime but not in the past 12 months) | 6.1% ± 0.5% | 8.5% ± 0.6% | 6.7% ± 0.5% | 7.7% ± 0.8% | 10.9% ± 0.7% |  |
| Former users, recent quitters (used in the past 12 months but not in the past 30 days) | 3.3% ± 0.3% | 4.5% ± 0.4% | 3.7% ± 0.3% | 5.5% ± 0.8% | 6.3% ± 0.7% |  |
| Current users (used in the past 30 days) | 4.1% ± 0.5% | 7.9% ± 0.8% | 7.7% ± 0.8% | 16.4% ± 1.5% | 24.9% ± 2.4% |  |
| Incomplete information or refused to answer | 3.3% ± 0.4% | 2.9% ± 0.3% | 2.3% ± 0.3% | 3.0% ± 0.7% | 5.9% ± 0.6% |  |
| **Among recent quitters and current users: Most common source of e-cigarette in the past 12 months** | (n=511) | (n=796) | (n=694) | (n=453) | (n=806) |  |
| Own purchase | 36.9% ± 2.4% | 45.9% ± 2.0% | 48.3% ± 2.2% | 53.1% ± 3.4% | 47.1% ± 1.5% | **0.007** |
| Nominee purchase (buyer age more than 20 years) | 5.0% ± 0.9% | 3.0% ± 0.7% | 4.6% ± 0.8% | 6.8% ± 1.5% | 2.8% ± 0.6% |  |
| Nominee purchase (buyer age less than 20 years) | 6.5% ± 1% | 3.9% ± 0.7% | 3.0% ± 0.7% | 2.6% ± 0.8% | 1.2% ± 0.4% |  |
| Given by others (buyer age more than 20 years) | 3.4% ± 0.8% | 4.9% ± 1.0% | 4.0% ± 0.9% | 3.7% ± 1.0% | 4.3% ± 0.9% |  |
| Given by others (buyer age less than 20 years) | 8.0% ± 1.5% | 5.3% ± 1.0% | 5.9% ± 1.1% | 4.8% ± 1.0% | 5.6% ± 1.0% |  |
| Stolen | 0.0% ± 0.0% | 0.7% ± 0.3% | 0.9% ± 0.4% | 0.9% ± 0.4% | 1.7% ± 0.5% |  |
| Others | 11.9% ± 1.9% | 6.8% ± 1.2% | 8.3% ± 1.1% | 7.1% ± 1.5% | 8.9% ± 1.5% |  |
| Refused to answer | 28.3% ± 2.3% | 29.5% ± 1.9% | 24.9% ± 2% | 21.0% ± 2.0% | 28.4% ± 2.1% |  |
| **Among recent quitters and current users: Type of e-cigarette used in the past 12 months** | (n=511) | (n=796) | (n=694) | (n=453) | (n=806) |  |
| Type 1 (looking similar to cigarettes) | 4.6% ±0.9% | 7.6% ±1% | 5.5% ±0.9% | 5.2% ±0.9% | 7.1% ±1.1% | **0.011** |
| Type 2 (pen) | 0.2% ±0.2% | 0.6% ±0.3% | 0.6% ±0.3% | 0.4% ±0.3% | 0.5% ±0.2% |  |
| Type 3 (tank) | 5.5% ±1% | 3.3% ±0.6% | 4.0% ±0.7% | 4.7% ±1.2% | 6.1% ±1.2% |  |
| Type 4 (electric pods) | 68.6% ±2.2% | 68.2% ±2.2% | 70.0% ±2.0% | 64.0% ±3.8% | 56.0% ±2.9% |  |
| Type 5 (toy-like pods) | 8.8% ±1.6% | 8.1% ±1.1% | 7.6% ±1.0% | 13% ±2.1% | 12.1% ±1.8% |  |
| Others | 2.2% ±0.7% | 2.5% ±0.5% | 3.1% ±0.9% | 2.6% ±0.9% | 6.0% ±1.2% |  |
| Refused to answer | 10.0% ±1.5% | 9.7% ±1.3% | 9.3% ±1.2% | 10.0% ±1.8% | 12.2% ±1.5% |  |
| **E-cigarette addiction screening test results (among current users only)** | (n=282) | (n=511) | (n=470) | (n=342) | (n=642) |  |
| Not addicted (0-3 points) | 26.4% ± 2.7% | 24.3% ± 1.5% | 28.2% ± 3.2% | 23.7% ± 2.4% | 23.0% ± 1.9% | 0.399 |
| Mildly addicted (4-8 points) | 27.3% ± 2.8% | 30.1% ± 2.1% | 27.3% ± 2.5% | 31.1% ± 3% | 32% ± 2.5% |  |
| Moderately addicted (9-12 points) | 16.4% ± 2.2% | 18.6% ± 1.8% | 18.1% ± 2% | 20.3% ± 2.8% | 17.6% ± 1.1% |  |
| Severely addicted (13 points) | 10.6% ± 2.2% | 13.4% ± 1.4% | 15.8% ± 2% | 11.6% ± 2% | 12.1% ± 1.4% |  |
| Incomplete information or refused to answer | 19.3% ± 2.9% | 13.5% ± 2.1% | 10.6% ± 1.7% | 13.4% ± 2.9% | 15.4% ± 1.9% |  |

**Remarks:** VC2 Commercial = Vocational Certificate 2 (Year 11, vocational) - Commercial track; VC2 Technical = Vocational Certificate 2 (Year 11, vocational) - Industrial technology track

## DISCUSSION

In this study, we described e-cigarette use behaviors among school-going adolescents in Thailand, disaggregated by sex assigned at birth and level of study. We found that one-quarter of Thai adolescents had used e-cigarettes in their lifetime, that the most common source of e-cigarettes was by the participants’ own purchase, that electric pods were the most common type of e-cigarettes used by the participants, and that one-third of all current users had moderate to severe levels of e-cigarette addiction symptoms. There were relatively small variations by sex compared to variations by level of study and educational system. The findings of this study should be of interest to stakeholders in tobacco control and adolescent health.

We provided more insights than previous analyses of nationally-representative data(Wichaidit et al., 2023, 2024) as we looked into additional details regarding e-cigarette use (e.g., sources, type of device). However, considering that dosage is a function of frequency, intensity, and duration(Wichaidit et al., 2019), future studies should consider how to measure the use of e-cigarettes in a more comprehensive manner. For use of smoking cigarettes, pack-years is the unit commonly used for measurement of both current and past cigarette use, with the common assumption that 20 cigarettes equal one pack(Kang et al., 2025). In e-cigarette use, issues with such calculation include the lack of a standard definition of one unit of use. We used the phrase "*15 puffs or 10 minutes*" in our study, as part of the e-cigarette addiction screening test, with an uncertain level of imprecision and error. Such a measurement primarily captures the frequency of e-cigarette use. The intensity of use depends on the content of the liquid vaporized by the e-cigarette device, which was not present on our questionnaire. The duration of use could not be estimated beyond the difference between the age at initiation and the age at the time of the Survey, which would be prone to over-counting. Participants might have years between initiation and the Survey when there was no e-cigarette use. Future studies that wish to focus on e-cigarette use should consider incorporating standard questions from larger studies such as the Global Adult Tobacco Survey (GATS)(Global Adult Tobacco Survey Collaborative Group, 2020) to gain additional insights.

Surprisingly, the prevalence of lifetime use of e-cigarette was nearly equal between male and female students. Such a similarity differed from the findings regarding other risk behaviors, such as alcohol consumption and cigarette smoking(Wichaidit et al., 2023), and may reflect the closing of behavior gaps between male and female students. Future studies should investigate this issue at greater depths, and also explore disparities by gender identities and sexual orientations. On the other hand, there were clear gaps between general education and vocational certificate students, even among those in the same year level (Year 11, i.e., Matthayom 5 vs. Vocational Certificate 2). The gaps could be explained, at least in part, by potential differences in health literacy and risk behaviors between the general education and vocational education systems(Orosram et al., 2026), and the issue deserves further in-depth investigations in future studies.

The strength of this study was the sample size and the sampling scheme, which enables the findings to be generalizable to adolescents from all regions of Thailand enrolled in formal education. However, our study also had a number of limitations. Firstly, although the investigators had taken measures aimed to reducing information bias, data collection nonetheless took place in the students’ own classroom during a school day on the use of a substance that was illegal according to local laws. Thus, the influence of social desirability could not be precluded from the study findings. Secondly, key details regarding e-cigarette use (e.g., brand, type of liquid placed in the device) were not included in this study to avoid excessive length, and should be considered in future studies that have a more specific focus on e-cigarette use. Thirdly, considering that up to 25 percent of Thai youths do not enrol in upper secondary education(Office of the Education Council, 2021) and we only collected data from school-attending students, the findings of the study may not be generalizable to a considerable proportion of adolescents who were outside of the formal education system. These limitations should be considered in the interpretation of the study findings.

## CONCLUSION

In this nationally-representative school-based cross-sectional study, we described the e-cigarette use behaviors of Thai adolescents in Year 7, 9, and 11 of a 12-year education system. Approximately one-quarter of Thai adolescents had used e-cigarette in their lives, and the most common source was by their own purchase. We found little variations by sex. However, vocational students were more likely than general education students to have used e-cigarettes. The findings of this study provide potentially useful basic information for stakeholders in tobacco control and adolescent health. However, limitations regarding potential biases, lack of important details, and the inability to generalize to adolescents not in the formal education systems should be considered as caveats in the interpretation of the study findings.

## Declaration of AI use

CH used ChatGPT to draft parts of the Introduction section to refine the sentences. CH also used ChatGPT in the Methods section to help refine the content based on the available protocols document. The details of both sections were then checked manually by WW before finalization with no additional generative AI input.

WW did not use any generative AI in the writing of the content of this manuscript. However, WW used Claude Opus 5.5 Medium to convert references for a part of the manuscript (provided by CH) to a Zotero library in RIS format. WW also had Grammarly installed on their desktop which automatically provided spell checks.

## Data Availability

All data produced in the present study are available upon reasonable request to the authors

